# Could Anti-retroviral treatment affects the profile of hepatitis B vaccine specific antibody in vertically HIV-infected children?

**DOI:** 10.1101/2023.09.22.23292573

**Authors:** T.F. Tchouangueu, L.B.M. Kouitcheu, A. Lissom, S.B. Tchuandom, J.C. Tchadji, C.S. Sake, G. Ambada, L. Ngu, H.F. Ouambo, C.O. Esimone, C.G. Chae Gyu Park, W.A. Alain Bopda, G.W. Nchinda

## Abstract

Paediatric immunisation had been relevant in reducing the widespread of Hepatitis B virus, as an outcome of the induction of hepatitis B surface antigen specific-IgG antibodies (anti-HBs). Studies revealed alteration effects of memory B cells during antiretroviral therapy (ART). We aimed at assessing anti-HBs response profile with respect to the most prominently used ART regimens in children.

a cross-sectional study was conducted in 116 participants made up of 72 HIV-exposed and infected children, subdivided into 20 antiretroviral-naïve on one hand and on another hand 52 ARV treated children made up of regimen subgroups, including 8 ABC-3TC-EFV/NVP (ART-R1), 19 ABC-3TC-LPV/r (ART-R2), 21 AZT-3TC-NVP (ART-R3) and 4 AZT-3TC-LPV/r (ART-R4), and 44 HIV-uninfected and unexposed (HUx or control group) children. Participants included in this study were regularly vaccinated children aged between 4 months and 5 years old, born to HIV-infected mothers. An optimized and adapted home-made ELISA and BioELISA® Biohit kit were used to measure specific IgM, IgG and IgG subclasses to HBs in children.

As result, this study showed that the rates of vaccine protective response in children treated with ART under regimens R1, R2, R3 and R4 were 25%, 38%, 51% and 75%, respectively. These protective response rates were significantly lower (p<0.0001) in children under R1, R2 and R3 than the control group (92%). When comparing anti-HBs specific IgM and IgG response medians; IgM response levels were similar in both control and ARV treated children, whereas R1 (p=0.0045), R2 (p=0.0016), and R4 (p<0.0001) showed significantly lower IgG level compared to the control group. Anti-HBs IgG subclass profile pattern in the control was IgG3≈IgG1≈IgG4>IgG2. However, IgG3≈IgG1≈IgG4>IgG4 profile pattern was estimated for children submitted to R1, R2 and R4, and the profile pattern of IgG3>IgG1≈IgG4≈IgG2 in those treated with R3 which also showed the most prominent anti-HBs IgG response mean rank level.

## 1. Introduction

About 90% of human immunodeficiency virus (HIV) and hepatitis B virus (HBV) infections in children are vertically acquired from mother-to-child transmission. Since these two viruses have similar transmission routes including sexual and percutaneous contacts, chronic HBV infection are commonly diagnosed in persons living with HIV (PLHIV) [1]. Almost 10% of the HIV-infected population are children and nearly all (95%) acquired HIV in childhood. Improvement in the HIV/AIDS management through routine detection, closed follow up and provision of highly active antiretroviral therapy (HAART) and prevention of mother-to-child transmission strategies has reduced the incidence of HIV during these recent years. Antiretroviral therapy (ART) had been among the most important interventions that resulted in the reduced burden of HIV.

Drug therapy and regimen characteristics can have a significant impact on the achievement of specific treatment goals. Advances in ART have made HIV a chronic disease that can be managed with lifelong drug therapy[2]. Controlling viral load, restoring immune function and preventing long-term complications are essential for successful HIV management, but some ARVs have been reported to be toxic. In fact ART-associated adverse effects vary from acute and potentially life threatening to chronic and insidious. These include hypersensitivity reactions (after Abacavir intake) and other life-threatening events such as symptomatic hepatotoxicity and severe cutaneous reactions [3]. Urolithiasis and renal tubulopathy caused by tenofovir disoproxil fumarate (TDF) and atazanavir respectively, and dyslipidemia account for toxicities that are not life threatening [3–8].

The use of ARV contributed to the control of HIV, but the most effective strategy in the case of HBV is through prevention, as this is essential in reducing its spread by vaccinating especially during childhood. In the absence of vaccination, there is increased risk of chronicity (> 90%) in infants who are exposed and infected to HBV early in their life. Hepatitis B virus infection is a global health concern, with a worldwide estimate of more than two billion individuals infected at least once in their lives [9, 10]. Its burden greatly depends on infections acquired in the first years of life[11]. In Sub-Saharan Africa the majority of the infections occur prenatally or during early childhood [12] and hence can easily progress to become chronic (with a rate >90%) in adulthood [13]. This is further compounded by concurrent HBV-HIV infection as it escalates the chronic HBV infection rate and also accelerates liver disease progression [14]. In addition, increase HBV replication rates combined with unusual persistence of HBV envelope antigen (HBeAg) was reported in HIV positive individuals, these are involved in relatively higher HBV pathology and transmission risk [15].

Protection against HBV is guaranteed at over 90% in immune competent children and adults after administration of the standard three dose HBV immunization schedules [16]. Protective HBV specific immunity is defined as the plasmatic concentration of HBsAg specific antibodies greater or equal to 10 mUI/ml[17, 18]. Many studies discussed the reduced hepatitis B vaccine response rate in HIV infected adults ranging from 46.9% and 85% [19–25]. This lower anti-HBs concentrations are associated with an overall rapid depressed response over time [19, 21, 26, 27]. To ameliorate this immune response in HIV infected persons, many vaccination strategies had been assessed to determine the optimal hepatitis B vaccine regimen and therefore heighten persistent response. These include increasing the number of injections or doses, the use of alternative routes of administration, or the use of adjuvants, and the newly proposed regimens are diverse and varied from three double-doses (40 mg at months 0, 1, and 6) [21, 23], four double-doses (40 mg at months 0, 1, 2, and 6) [19, 20, 22, 24] to four standard-doses (20 mg at months 0, 1, 2, and 6) [22, 24, 25]. Hence, anti-HBs protective plasmatic rate reduced from 71% at year 1 to 40% by year 5 after three doses of 40 mg of hepatitis B vaccine in HIV infected adults [27].

While these strategies could potentially increase HBV vaccine specific antibody titers in HIV infected individuals little is known about the profile of anti-HBs specific IgG subclass responses associated with the vaccination of HIV-infected children. Thus, assessing anti-HBs IgG subclass responses following HBV vaccination of HIV infected children could provide important data pertaining to the maturation and functions of the vaccine-induced humoral immune responses with respect to ART. Following natural seroconversion, IgG subclass responses specific to HBsAg have been demonstrated to be restricted mainly to neutralizing IgG1 and IgG3, with only a minor contribution from IgG2 and IgG4 [28, 29]. The presence of both IgG1 and IgG3 is strongly associated with Th1 cytokines such as IL-2, IFN-γ and TNF-α which are critical in cell-mediated immunity and the induction of memory IgG responses [30]. Activation of Th1 response together with HBsAg specific IgG1 or IgG3 subclass responses is thus a critical necessity in the ultimate effective neutralization of HBV [31]. However, there is limited data about the profiles of HBsAg specific IgG subclass responses of HIV-1 infected children following childhood HBV vaccination [32, 33]. The aim of this study was to assess the profiles of HBsAg specific IgG subclass responses in ARV-treated HIV infected children.

## 2. Methods and Participants

### 2.1. Study design

We conducted a cross-sectional multicentric study during a period of 14 months from December 2014 to March 2016 in five health centres in the surroundings of Yaounde, including the Yaounde University Teaching Hospital (CHUY), Efoulan District Hospital (EDH), Bikop Catholic Health center (CSCB), Social and Health Animation Center (CASS); and the Chantal Biya International Reference Centre for Research on the Prevention and Management of HIV/AIDS (CIRCB).

The study participants were selected using consecutive sampling technique and written informed consent was obtained from parents or legal guardians of the children study participants. The inclusion criteria for the enrolment of children in this study was Children aged 4 months to 5 years who had completed the pediatric HBV vaccination according to the national program on immunization. Pediatric HBV vaccination was confirmed through vaccination records. We excluded all children whose parents or legal guardians refused to sing a written consent form, those who provided incomplete data and those with incomplete pediatric HBV vaccination. We equally excluded children who were positive to prevalent endemic infections including HBV, HCV, Dengue virus infection and Malaria, after a screening.

### 2.2. Sample collection and processing

Blood collection was done at least 4 weeks after the last HBV vaccine dose. About three milliliters of peripheral blood were collected from each participant in Ethylene Diamine Tetra-Acetic Acid (EDTA)-containing tubes. All samples were stored at room temperature and processed within 2 hours of collection. To obtain plasma, samples were centrifuged at 2,000 rpm for 10 min at 4°C. The plasma fraction was harvested under sterile conditions in a level II biosafety cabinet, aliquot in small, single-use volumes and stored at −20°C until use.

The screening of malaria, dengue, hepatitis B and hepatitis C virus infections was performed in all samples. The malaria diagnostic and CD4/CD8 cell counts were done in whole blood, while plasma samples were used for all serological analysis. Malaria diagnosis was done using an SD BIOLINE® point of care kit (Giheung, Republic of Korea). SD BIOLINE® HBsAg and anti-HCV immuno chromatographic tests were used for the diagnosis of HBV and HCV, respectively. The CTK®OnSite (San Diego, USA) Duo Dengue Ag-IgG/IgM rapid test was used for the simultaneous detection and differentiation of DENV specific IgM and IgG antibodies as well as NS1 antigen for all the samples. Absolute numbers of CD4/CD8 cells for HIV-1^+^ participants were determined using BD multi test CD3/CD8/CD45/CD4 and TruCount tubes (BD Biosciences, San Jose, USA) according to the manufacturer’s instructions.

### 2.3. Quantification of plasma levels of anti-HBs antibodies

Quantification of Anti-HBs antibody in participants’ plasma was done using BioELISA anti-HBs Kit (Biokit S.A, Barcellona, Spain) according to the manufacturer’s instructions. Briefly, 100μl/well of each participant’s plasma was added in an ELISA plates pre-coated with highly purified HBsAg. After which plates were incubated at 37°C for one hour before washing for 4 times with diluted washing buffer solution provided with the kit. The Horseradish Peroxidase (HRP)-conjugated anti-human IgG antibodies were added (100 μl/well) and the plates were incubated at 37°C for 30 minutes. Then after four times washing, 100μl/well of substrate was then added and incubated at room temperature for 30 minutes. The reaction was stopped with 50 μl/well of sulphuric acid (stop solution) and the optical densities (OD) determined at 450 nm. The concentration of anti-HBs for each participant was calculated from the calibration curve generated from provided pre-diluted standards according to the manufacturer’s instructions. Anti-HBs concentration in this study were categorized as not reactive (< 10 mIU/ml), and reactive and protective (≥ 10 mIU/ml).

### 2.4. Quantitative determination of anti-HBs antibodies

Plasma samples were analysed for anti-HBs specific antibody responses using an optimized in-house ELISA protocol previously described [32]. Briefly 96-wells flat-bottomed high binding Costar® assay plates (CORNING, USA) were coated either with recombinant HBsAg (IMMUNODX Woburn, MA, USA) dissolved in PBS (50 ng/well) and incubated at 4°C overnight.

The following day, plates were washed three times with PBST (PBS with 0.05 % Tween-20) and blocked with 3% BSA (Carl ROTH, Karlsruhe, Germany) for one hour at 37°C. After an additional washing step, plasma diluted (1:500) in PBS was added (100 µl/well) in the corresponding wells in triplicates and incubated for two hours at 37°C. The plates were washed five times as described above, then 100µl HRP-conjugated anti-human IgG (1:2000), HRP-conjugated anti-human IgG1, IgG2, IgG3, IgG4, or IgM (1:4000) antibodies were added in the appropriate plates and incubated for 1h at 37°C. The secondary antibodies used in this study were mouse anti-human IgG Fc (clone JDC-10), mouse anti-human IgM (clone UHB), mouse anti-human IgG1 Fc (clone HP6001), mouse anti-human IgG2 Fc (clone 31-7-4), mouse anti-human IgG3 Hinge (clone HP6050) and mouse anti-human IgG4 pFc’ (clone HP6023), all HRP conjugated antibodies were procured from Southern Biotech® (Birmingham, USA). Plates were washed five times and 100µl of 2,2′-Azino-bis (3-ethylbenzthiazoline-6-sulfonic acid) substrate was added to each well and incubated in the dark for 30 min at 37°C. The enzyme-substrate reaction was stopped by adding a stop solution (Southern Biotech, Birmingham, USA). The optical densities (OD) at 405nm were read using a multiscan Fc Elisa microplate reader (Thermoscientific, USA). The mean OD values were determined after normalization with background values derived from wells treated with validated anti-HBs plasma samples similarly treated.

Dilution of HBs antigen, plasma samples and secondary antibodies were previously separately optimize, and a decision was held for a single dilution to each. Relatively increased plasmatic levels of anti-HBc antibodies, concomitantly tested in every samples, was considered as an indicator of HBV exposure and infection of these children. Hence anti-HBs antibodies obtained in their blood will be a result of previous exposure to the virus during neonatal or maternal infections rather than the course of vaccine immune response.

### 2.6. Statistical analysis

Continuous variables from children’s characteristics and antibody response profiles were described as medians and Inter Quartile Ranges (IQR) and categorical variables were presented as percentages. Comparison between groups were made using the non-parametric Mann-Whitney U test for continuous variables and Chi square (or Fischer exact) tests for categorical variables as appropriate. The level of statistical significance was set at p < 0.05. Statistical analysis was performed using Graph Pad Prism version 9.0.0 software.

## 3. Results

### 3.1. Study Population Characteristics

The study characteristics presented in **Table 1**, included 116 participants, amongst who 72 were HIV-1 exposed infected (HEI) children with by their mother, whereby 52 of them were treated with ART (HEI/ARV+) and 20 were not (HEI/ARV-). The median ages amongst these groups were 35.25 (IQR 25.12 – 49.00) and 25.75 (IQR 12.50 – 40.50) months, respectively. A total 26 and 9 females with of 32.0 and 27.0 months, as well, 26 and 11 males with median ages of 38.5 and 24.5 months (IQR 34-59) in HEI children following ARV (ARV+) or not (ARV-), respectively, were recruited. The median (IQR) CD4+ cells count in children were 1992 (1317-2702) cells/mm^3^ respectively in HIV/ARV+ and 1602 (1011-3055) and 2004 (1179-2885) cells/mm^3^ in HEI/ARV+ and HEI/ARV-, respectively. As controls, 44 HIV negative children with an overall median age of 11.5 months (IQR, 7-23.50) were also recruited. All participants completed the three-dose HBV vaccination schedule and were all negative to HBsAg and HBV envelope antigen (HBeAg) as well as anti-HBc antibodies.

**Table 1:** Study Population Characteristics.

| Variables |  | HUx |  | HEI/ARV- |  | HEI/ARV+ |  |
| --- | --- | --- | --- | --- | --- | --- | --- |
|  |  | Female | Male | Female | Male | Female | Male |
| Participant n (%) |  | 20 (45.5) | 24 (54.5) | 26 (50.0) | 26 (50.0) | 9 (45.0) | 11 (55.0) |
| Median age, [IQR],<br>(Range) in months (a) |  | 10.0<br>[7-23]<br>(4-59) | 13.0<br>[7-24]<br>(5-59) | 32.0<br>[18-49]<br>(5-60) | 38.5<br>[32.25-49]<br>(6-60) | 27.0<br>[11.5-38]<br>(10-59) | 24.5<br>[13.5-43]<br>(7-60) |
| Median CD4 Count,<br>[IQR],<br>(Range) in cells.mL-1(a) |  | N/A | N/A | 1696<br>[1012-3129]<br>(335-4932) | 1508<br>[1009-2982]<br>(534-4283) | 2015<br>[1041-3062]<br>(1035-5315) | 1992<br>[1317-2702]<br>(1085-4478) |
| Mean ±SD of CD4/CD8<br>Ratio(b) |  | N/A | N/A | 2.56 ± 0.96 | 1.91 ± 1.07 | 1.27 ± 0.57 | 1.05 ± 0.69 |
| Feeding<br>options<br>n (%)<br>Mixed | Breastfeeding | N/A | N/A | 13 (50.00) | 8 (30.77) | 4 (44.44) | 4 (36.36) |
|  | Mixed | N/A | N/A | 3 (11.54) | 9 (26.92) | 3 (33.33) | 3 (27.27) |
|  | Formula | N/A | N/A | 10 (38.46) | 7 (37) | 2 (22.22) | 4 (36.36) |

ART: Antiretroviral therapy; IQR: Interquartile range; n: number; (a): Mann-Whitney test comparing median age of a given gender between groups. (b): unpaired t test comparing mean CD4/CD8 ratios between genders of a group; HUx: HIV-1 unexposed and uninfected children; HEI: HIV-1 exposed and uninfected children; HEU: HIV-1 exposed and uninfected children; PMTCT: prevent mother-to-child transmission; N/A: Not Applicable

### 3.2. Seroprotective response of antibody to paediatric Hepatitis B virus surface antigen vaccine

The seroprotective response rate of paediatric HBV vaccine in both ART (41%) and ART naïve (50%) HIV-infected children (HEI/ART-and HEI/ART+) were significantly lower than in their control (HUx 93%) peers (p=0.0384 and p=0.0027, respectively) (**Figure 1A**). In addition, similar anti-HBs IgM response was observed in the three groups of participants. The anti-HBs IgG specific response levels in both HEI/ART- and HEI/ART+ children were significantly reduced (p<0.0001) compared to their control peers (**Figure 1B**).

**Figure 1:**
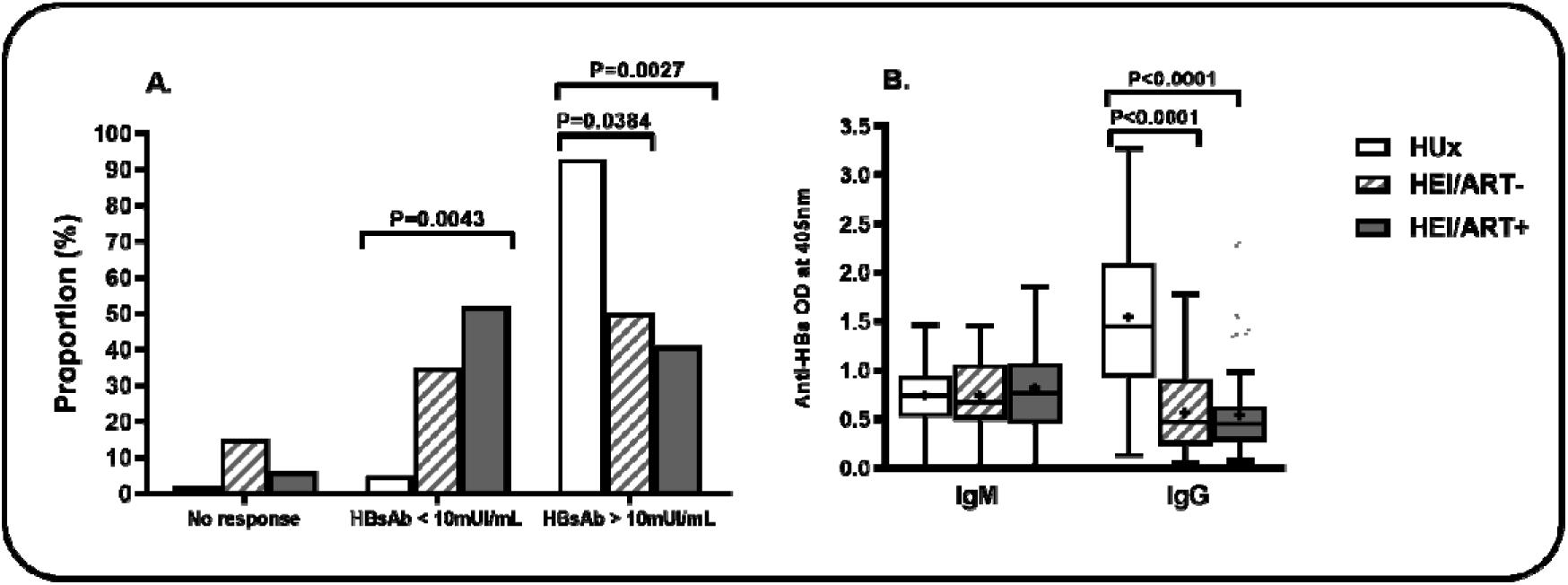
Hepatitis B surface antigen vaccine response rate and anti-HBs specific antibody responses according to post-natal ART in HIV infected children. In (A), is shown the proportion of children who responded to HBV vaccine in HIV vertically infected children depending on ART; (B), The anti-HBs specific IgM and IgG response in HIV infected children. HUx: HIV unexposed and uninfected children, HEI/ART-: antiretroviral naïve HIV exposed infected children; HEI/ART+: antiretroviral treated HIV exposed infected children. Statistical analyses were perfomed using Chi square test for goodness of fit to compare response rates for each of the response rates patterns (A) and Kruskal-Wallis test with Dunns’multiple test for antibodies response levels amongst IgM and IgG antibodies (B). p-value is considered significant at p<0.05.

Moreover these response rates were highly variable depending on the ART regimen (ARV/R). As shown in figure 2, hepatitis B vaccine protective response in children treated with ART under regimens R1, R2, R3 and R4 were 25%, 38%, 51% and 75% respectively. The proportions of HBV vaccine protected children and treated with regimens R1, R2 and R3 were significantly lower than the control (p<0.0001 for each of them), and also than those treated with ARV R4 (p<0.0001, p=0.0006 and p=0.004, respectively) (**Figure 2A**). Nevertheless, non-protective HBV vaccine responses were shown in high proportions of children treated with ARV regimens R1 (75%), R2 (61%) and R3 (40%). As commonly seen, anti-HBs-specific IgM levels in both control and ARV treated children were similar irrespective of the treatment regimen. The anti-HBs specific vaccine response mean ranks in ARV-treated children with regimens such as R1 (p=0.0045), R2 (p=0.0016), and R4 (p<0.0001) were significantly lower than that of children in the control group (**Figure 2B)**

**Figure 2:**
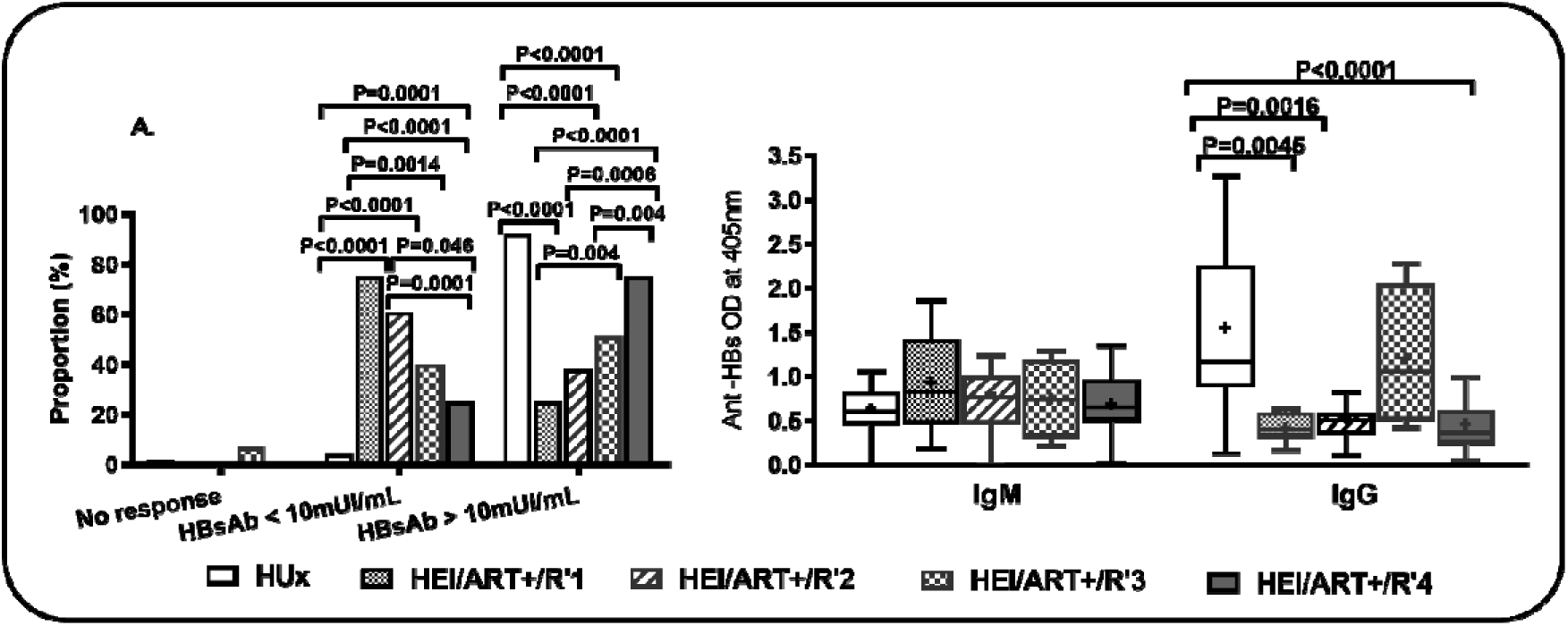
Hepatitis B surface antigen specific antibody responses rate and levels in vertically infected regarding ARV regimens. In (A), is shown the proportion of children who responded to HBV vaccine in both HIV infected children with respect ART; In (B), The anti-HBs specific IgM and IgG response in HIV infected children with respect to ART regimens. HUx: HIV unexposed and uninfected children, HEI/ART-: antiretroviral naïve HIV exposed infected children; HEI/ART+: antiretroviral treated HIV exposed infected children. R1:ABC-3TC-EFV/NVP, R2:ABC-3TC-LPV/r, R3: AZT-3TC-NVP and R4: AZT-3TC-LPV/r. Statistical analyses were perfomed using Chi square test for goodness of fit to compare response rates for each of the response rates patterns (A) and Kruskal-Wallis test with Dunns’multiple test for antibodies response levels amongst IgM and IgG antibodies (B). p-value is considered significant at p<0.05.

### 3.3. Hepatitis B vaccine specific IgG subclass responses with regard to post-natal ART in HIV infected children

A closer look into the response of IgG response was observed with the four subclasses as underlined in **Figure 3**. In antiretroviral naïve children, anti-HBs specific IgG1, IgG2, IgG3 and IgG4 were all significantly lower (p=0.0113, p=0.0007, p=0.0031 and p=0.0003, respectively) compared to the control (**Figure 3A**). The same pattern was observed with the antiretroviral treated participants p<0.0001, p<0.0001, p=0.0008 and p<0.0001, respectively). We noticed as far as the IgG profile response pattern is concerned that similar profile patterns in both control (HUx) and HEI/ART+ as IgG3≈IgG1≈IgG4>IgG2 (**Figure 3B and 3D**). A relatively different pattern of IgG1≈IgG3≈IgG4≈IgG2 was specific to HEI/ART-children (**Figure 3C**)

**Figure 3:**
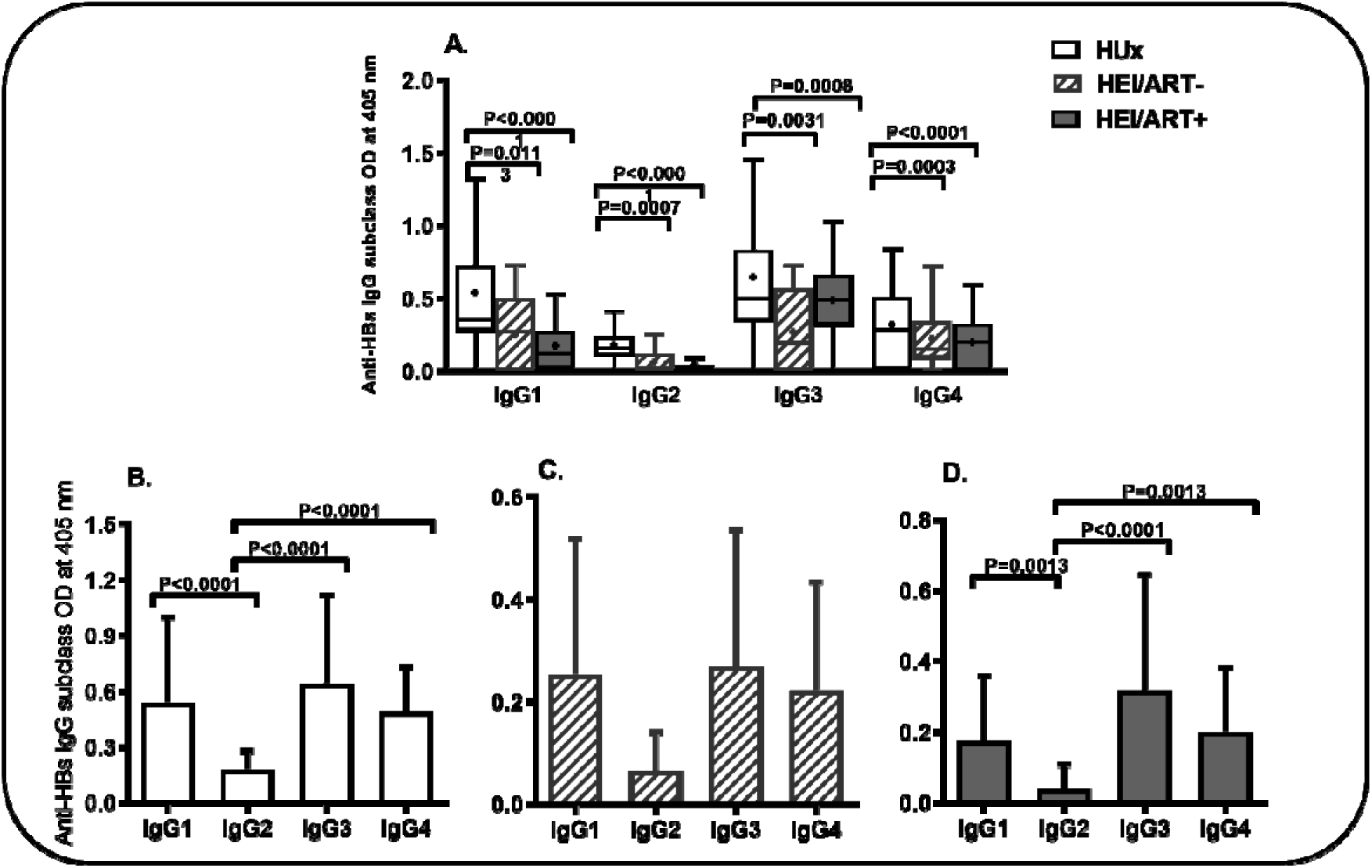
Hepatitis B surface antigen specific IgG subclass responses in HIV vertically infected children regarding ART. In (A), is shown the anti-HBs specific IgG subclass response in HEI children regarding ART. Profile of Hepatitis B surface antigen specific IgG subclass responses in HUx (B), HEI/ART-(C) and HEI/ART+ (D). HUx: HIV unexposed and uninfected children, HEI/ART-: antiretroviral naïve HIV exposed infected children; HEI/ART+: antiretroviral treated HIV exposed infected children. (B), (C), and (D) graphs were used to determine the pattern of IgG subclass responses with respect to each of the group of participants. Statistical analyses were perfomed using Kruskal-Wallis test followed by Dunn’s multiple comparison test to compare IgG antibody subclass response levels between groups of children (A) then for each of the groups HUx (B), HEI/ART-(C), and HEI/ART+ (D) children to compare IgG subclass response levels. p-value is considered significant at p<0.05

There were poor variations between ARV treated HIV infected groups of children and their control peers in terms of anti-HBs-specific IgG subclass response levels, though a significantly lower anti-HBs specific IgG1 and IgG4 responses was shown in children undergoing treatment with regimen R2 ARV compared to the HUx children (p<0.0001 and p=0.0012, respectively) (**Figure 4A**). Moreover, specific IgG4 response in ARV-treated children with R3 was lower (p=0.0012) than in the controls. Almost all the IgG subclass profile patterns were similar in ARV-treated pools of children and the HUx ones. IgG3 subclass remained the most expressed antibody for all subgroups. The profile IgG3≈IgG1≈IgG4>IgG2 (**Figure 4B**) was shown in the HUx children and relatively variable profile patterns were estimated for HEI/ART^+^/R1 (**Figure 4C**), HEI/ART^+^/R2 (**Figure 4D**) and HEI/ART^+^/R4 (**Figure 4E**) subset of children as IgG3≈IgG1≈IgG4>IgG2 against IgG3>IgG1≈IgG4≈IgG2 in HEI/ART^+^/R3 ones (**Figure 4F**).

**Figure 4:**
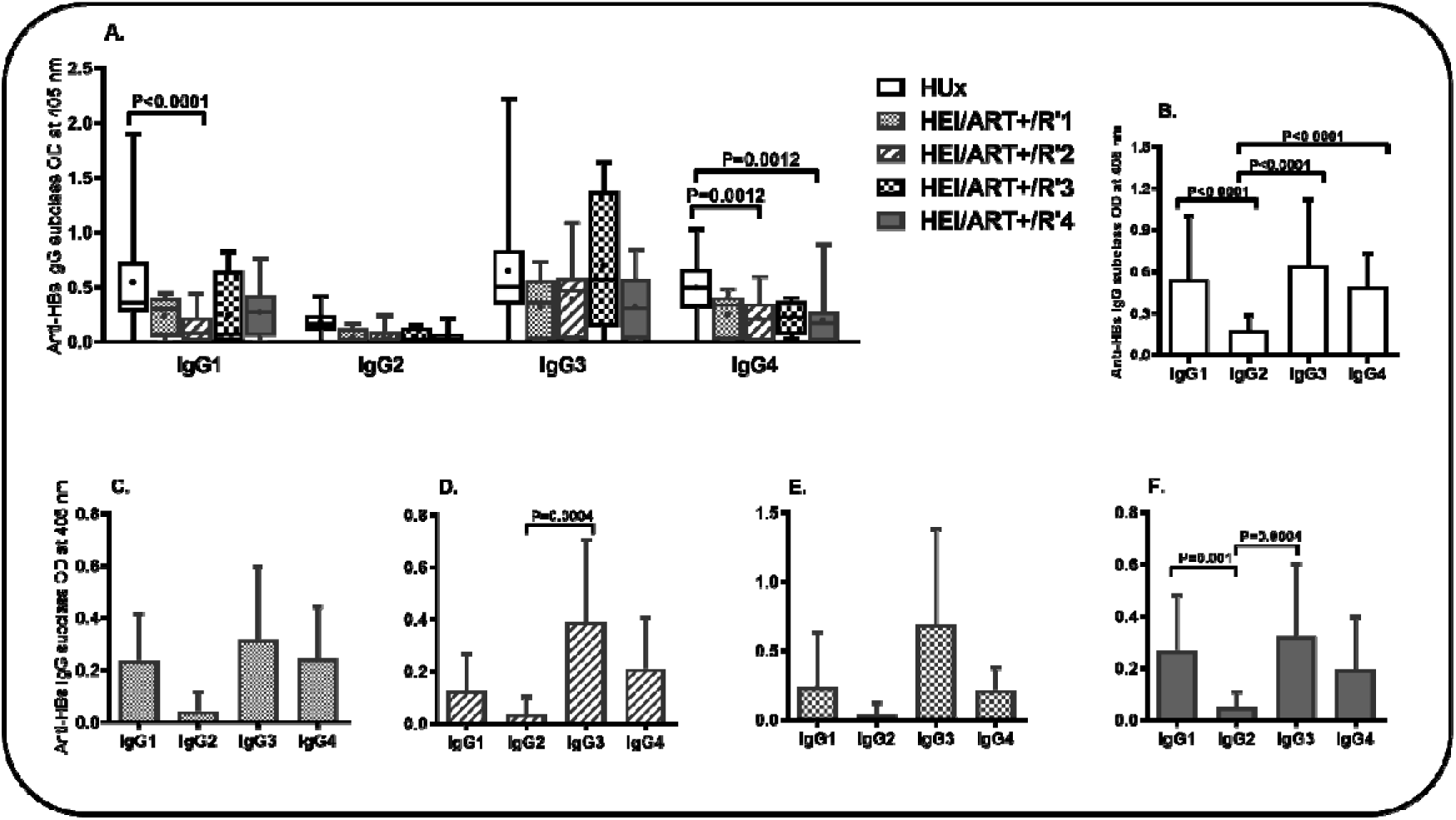
Hepatitis B surface antigen specific IgG subclass responses in HIV infected children regarding ART regimen. In (A), is shown the anti-HBs specific IgG subclass response in HEI children regarding ART regimen. Profile of Hepatitis B surface antigen specific IgG subclass responses in HUx (B), HEI/ART+/R1 (C), HEI/ART+/R2 (D), HEI/ART+/R3 (E) and HEI/ART+/R4 (F). HUx: HIV unexposed and uninfected children; HEI/ART+: antiretroviral treated HIV exposed infected children. R1:ABC-3TC-EFV/NVP, R2:ABC-3TC-LPV/r, R3: AZT-3TC-NVP and R4:AZT-3TC-LPV/r. (B), (C), (D) and (E) graphs were used to determine the pattern of IgG subclass responses with respect to each of the ART regigem intake by these participants. Statistical analyses were perfomed using Kruskal-Wallis test followed by Dunn’s multiple comparison test to compare IgG antibody subclass response levels between groups of children (A) then for each of HUx (B), HEI/PMTCT+/R1 (C), HEI/PMTCT+/R2 (D) HEI/PMTCT+/R3 (E) and HEI/PMTCT+/R4 (F) groups of children to compare IgG subclass response levels. p-value is considered significant at p<0.05.

### 3.4. Association between CD4/CD8 ratio and Hepatitis B vaccine specific IgM or IgG response with regards to post-natal ART

This study revealed that there was a negative independent correlation between CD4/CD8 ratio and anti-HBs specific IgM in both subgroups of children. Whereas, significant dependent slightly decreased association between IgG response and both HEI/ART-(r=-0.398, p=0.0196) and HEI/ART+ (r=-0.527, p=0.014) (**Table 2**). Then, a positive independent association between CD4/CD8 ratio and anti-HBs specific IgM response, as well as a negative non-significant association between CD4/CD8 and IgG response in HEI/ART^+^/R1 subgroup of children. However, no association was noticed in both HEI/ART^+^/R2 and HEI/ART^+^/R3 subgroup of children irrespective of the studied antibodies (**Table 3**).

**Table 2:** Correlation between CD4/CD8 ratio and Hepatitis B vaccine specific IgM or IgG response with regard to post-natal ART.

| Antibody isotypes | HEI/ART- |  | HEI/ART+ |  |
| --- | --- | --- | --- | --- |
|  | r | p-value | r | p-value |
| **IgM** | -0.166 | 0.364 | -0.206 | 0.572 |
| **IgG** | -0.398 | 0,0196* | -0.527 | 0.014* |

HEU: HIV exposed uninfected children, HEI: HIV exposed infected children, r: Spearman correlation coefficient; P: p-value at 0.05 significant level; \*: p<0.05.

**Table 3:** Correlations between CD4/CD8 ratio and anti-HBs IgM and IgG antibody responses.

| Antibody isotypes | HEI/ART<sup>+</sup>/R1 |  | HEI/ART<sup>+</sup>/R2 |  | HEI/ART<sup>+</sup>/R3 |  |
| --- | --- | --- | --- | --- | --- | --- |
|  | r | p-value | r | p-value | r | p-value |
| **IgM** | 0.4 | 0.75 | -0.071 | 0.802 | 0.038 | 0.906 |
| **IgG** | -0.6 | 0,417 | 0.028 | 0.923 | 0.043 | 0.892 |

vertically HIV infected children with regard to ARV regimen for therapy
HEI/ART+: antiretroviral treated HIV exposed infected children. R1: ABC-3TC-EFV/NVP, R2: ABC-3TC-LPV/r, R3: AZT-3TC-NVP. r: Spearman correlation coefficient; P: p-value at 0.05 significant level.

## 4. Discussion

Hepatitis B Virus infections are more likely to progress to chronic infections amongst children who acquire HBV at early stage of their lives [34]. In high HBV prevalence areas, such as sub-Saharan Africa (SSA) regions including Cameroon, the majority of HBV infections occur during the perinatal period or early childhood because of close household contact, medical or cultural procedures like scarification and tattoo [35]. In children without HBV immunization, the risk of chronicity is even greater (> 90%). Children in this study were completely negative to anti-HBc and HBsAg as this supports the evidence that low risk of HBV vertical transmission in such a typical tropical SSA setting [36, 37]. Hence, vaccine preventing HBV infection is essential for both HIV infected and uninfected persons and its specific antibodies are sustained into blood at higher concentrations for at least five years after the last dose as were revealed by our data (Figure S1).

We have shown in this study that hepatitis adequate vaccine immune responsiveness (HBsAg≥10mIU/ml) after a three-dose intake in both antiretroviral treated and naïve HIV infected children was reduced. Significantly lower seroprotective response rate of paediatric HBV vaccine in both ART (41%) and ART naïve (50%) HIV-infected groups of children (HEI/ART- and HEI/ART+) as compared with their control (HUx 93%) peers (p=0.0384 and p=0.0027, respectively) was found. Similarly, significantly reduced (20% vs 60.71%) immune response rate was determined among age-related HIV exposed and infected children from the same community [38] as well as in a study done in Botswana where HIV-positive infants mounted a significantly lower immune response to the HBV vaccine (74.5% vs 89.2%) [39]. These findings are consistent with our results and can be explained by a faster decline of vaccine titer after the initial response among HIV infected children [40]. Previous studies discussed the importance of HAART in restoring HBV vaccine immune response through the improvement of plasmatic immune cells levels, mainly CD4^+^ T cells [38, 41–43]. Since heightened HIV plasmatic viral load will contribute in destroying immune cells leading to impairment of the immune system to the extent that the vaccine immunity to hepatitis B become substantially reduced or inexistent [44, 45]. However we were unable to find hepatitis B vaccine immune restoration in the current study although an overall increase in CD4^+^ T cell was observed in HIV/ARV+ group of children.

Given that the overall vaccine responsiveness was lower in HIV infected infant under ART than in their control peers, these levels were specifically dependent upon ART regimen used for HIV infection control. Nevertheless, children treated with the tri-therapy “AZT-3TC-LPV/r” showed a relatively higher HBV vaccine response rate (75%). Our research proup previously delineate the impairment of HBV vaccine immune response in children after *in-utero* ARV exposure [33]. Several studies confirmed that B cells abnormalities including hypergammaglobulinaemia and HIV specific and non-specific B-cell responses were decreased by ART [46–48]. Moreover, memory B-cell responses specific to vaccine antigens were also reported to be preserved in vertically infected children who received early HAART [49]. Memory B cells play an important role in a persistent immune response to a specific antigen. They are characterized as long-lived, resting, antigen-experienced cells that are rapidly inducible after exposure to cognate antigen [50]. Studies have shown that although ART leads to decreased numbers of CD27^+^ activated B cells and plasmablasts [49, 51], the increase in the number of CD27^+^ resting memory B cells following treatment occurs slowly and remains incomplete [51–53].

In addition, similar IgG1 and IgG3 responses was noticed among children with R3 therapy compared with their unexposed peers. Such observations suggest that AZT-3TC-NVP (R3) regimen played an important role in improving IgG immune response to HBV vaccine response. Unlike increased difficulties to determime from tritherapy the exact source of response improvement, we suggest zidovudine to be the odd among these three molecules. However, there is lack of evidence-based reports regarding direct impact of specific antiretroviral regimens on humoral response

Similar anti-HBs IgM response was observed among the three groups of participants. The anti-HBs specific response levels in both HEI/ART- and HEI/ART+ children were significantly reduced (p<0.0001) compared to their control peers. Moreover these response rates were highly variable depending on the ART regimen (ARV/R). In Fact, hepatitis B vaccine protective response in children treated with ART under regimens R1, R2, R3 and R4 were 25%, 38%, 51% and 75% respectively. Proportions of HBV vaccine protected children treated with regimens R1, R2 and R3 were significantly lower than the control (p<0.0001 for each of them), and also than those treated with ARV R4 (p<0.0001, p=0.0006 and p=0.004, respectively)

Previous studies have described the significance of CD4/CD8 ratio as a predictor of HBV vaccine response [54]. Persistent inflammation in HIV positive individuals is involved in accelerated ageing, monitored with low CD4/CD8 ratio and good immuno-virological responses [55, 56]. Mussini et al described that normalization of CD4/CD8 ratio after HAART initiation correlated with a lower risk of non-AIDS related events [57]. Our findings showed a negative association (r=-0.3854, p=0.0244) between CD4/CD8 ratio and anti-HBs IgG response in HIV exposed children born to mothers who did not participate to PMTCT. Hence ARV treatment for preventing mother-to-child transmission is needed for vaccine seroconversion and immune recovery in infants born from HIV infected mother. There are many indications supporting the increase rate of B-cell death during ongoing viral replication and B cells loss [58–60]. Also, memory B-cell responses and serological memory against both T-cell-dependent and T-cell-independent antigens are decreased in HIV-infected individuals [61–64].

The incidence of HIV infection among HIV vertically exposed infants and children have dropped with the prevention mother-to-child transmission (PMTCT) programs. The Option B+ approach, in which all HIV-infected pregnant women initiate lifelong ART irrespective of clinical or immunologic status, represents a paradigm shift in the field of PMTCT. However, various abnormalities in infants have been reported to be related to maternal ART in the prevention of vertical HIV infection [65–72]. More is yet to understand about the role that plays *in-utero* antiretroviral therapy from PMTCT in repressing or boosting this response.

## 5. Conclusion

Overall, we can conclude that in addition to the significant reduced humoral immune response among seropositive children treated or not to ARV, there was relative waning of the neutralizing IgG3 subclass among ARV-naive children compared with the ARV-naive peers. We noted a significantly diminished HBV vaccine immune response rate in HEI/ART+ children for all studied ARV regimens except for AZT-3TC-LPV/r, meanwhile the mean rank level of anti-HBs level of tritherapy “AZT-3TC-NVP” was comparable that in their control peers. Given that anti-HBs titer is a strong correlate for predicting protection after HBV vaccination; HIV infected children should be tested regularly for anti-HBs concentration in order to give booster injection when titers are below 10mIU/mL.

## Author Contributions

Conceptualization, T.T.F., K.M.L.B. and N.W.G; methodology, T.T.F., L.A., T.B.S., T.J.C, N.LN, S.C.S. and O.H.F; data analysis, T.T.F., L.A., investigation, T.T.F., A.G and S.C.S.; writing—original draft preparation, T.T.F.; writing review and editing, K.M.L.B., E.C.O., P.C.G., BWA and A.G.; All authors have read and agreed to the published version of the manuscript.

## Institutional Review Board Statement

The study was conducted in accordance with the Declaration of Helsinki, and approved by the Cameroon National Ethical Committee for research on health sciences under protocol number 2014/07/474/CE/CNERSH/ SP for studies involving humans.

## Informed Consent Statement

All parents or legal guardians signed a written informed consent for the participation of their children. Children that tested positive for any of prevalent endemic diseases were excluded from the study and recommended to the paediatrician for clinical follow up.

## Definition of terms and abbreviations

3TC: Lamivudine
ABC: Abacavir
Anti-HBc: Antibody to hepatitis B core antigen
Anti-HBs: Antibody to hepatitis B surface antigen
ART: Antiretroviral therapy
ARV: Antiretroviral
ATV/r: Atazanavir/ritonavir
AZT: Zidovudine
EDTA: Ethylene Diamine Tetraacetique Acid
EFV: Efavirenz
HBcAg: Hepatitis B Core Antigen
HBsAg: Hepatitis B Surface Antigen
HBV: Hepatitis B Virus
HEI: (HIV-Exposed and infected children): HIV-positive children who are born to HIV-infected mother
HEI/ARV: (Antiretroviral-naïve HIV-Exposed and infected children): HIV-positive children who are born to HIV-infected mother
HEI/ARV: (HIV-Exposed and infected children treated to antiretroviral): HIV-positive children who are born to HIV-infected mother
HUx: (HIV-unexposed and uninfected children): control group of children born to HIV negative mothers.
LPV/r: Lopinavir/ritonavir
NVP: Nevirapine
R/ART-R: Specific Antiretroviral therapy Regimen [(**R1/ART-R1**: Abacavir-Lamivudine-Efevirenz or Niverapine (ABC-3TC-EFV/NVP), **R2/ART-R2**: Abacavir-Lamivudine-Lopinavir/ritonavir (ABC-3TC-LPV/r), **R3/ART-R3**: Zidovudine-Lamivudine-Niverapine (AZT-3TC-NVP), **R4/ART-R4**: Zidovudine-Lamivudine-Lopinavir/ritonavir (AZT-3TC-LPV/r)]

## References

1. Bigna, J. J., Nkeck, J. R., Ngouo, A., Nyaga, U. F., Noubiap, J. J. J. I., Disease and Health Hepatitis B virus and HIV coinfection among adults residing in Cameroon: A systematic review and meta-analysis of prevalence studies, Infection, Disease & Health 2018. 23, 170–178.

2. Lee, C., Sapasap, J., LaRochelle, J., Smith, R. O. and Badowski, M. E. Antiretroviral Therapy in Children and Adolescents: A Look Into Modern Single Tablet Regimens, J Pediatr Pharmacol Ther 2021. 26, 783–794.

3. Mounzer, K., Hsu, R., Fusco, J. S., Brunet, L., Henegar, C. E., Vannappagari, V., Stainsby, C. M., Shaefer, M. S., Ragone, L. and Fusco, G. P. HLA-B*57:01 screening and hypersensitivity reaction to abacavir between 1999 and 2016 in the OPERA(®) observational database: a cohort study, AIDS research and therapy 2019. 16, 1.

4. Motisi, M. A., Tamborino, A., Parigi, S., Galli, L., de Martino, M. and Chiappini, E. J. J. o. C. The use of antiviral drugs in children, 2022. 34, 73–86.

5. Seo, J. W., Kim, K., Jun, K. I., Kang, C. K., Moon, S. M., Song, K. H., Bang, J. H., Kim, E. S., Kim, H. B., Park, S. W., Kim, N. J., Choe, P. G., Park, W. B. and Oh, M. D. Recovery of Tenofovir-induced Nephrotoxicity following Switch from Tenofovir Disoproxil Fumarate to Tenofovir Alafenamide in Human Immunodeficiency Virus-Positive Patients, Infection & chemotherapy 2020. 52, 381–388.

6. Bossi, P., Colin, D., Bricaire, F. and Caumes, E. Hypersensitivity syndrome associated with efavirenz therapy, Clinical infectious diseases : an official publication of the Infectious Diseases Society of America 2000. 30, 227–8.

7. Kim, G. Y., Anderson, K. R., Davis, D. M. R., Hand, J. L. and Tollefson, M. M. Drug reaction with eosinophilia and systemic symptoms (DRESS) in the pediatric population: A systematic review of the literature, Journal of the American Academy of Dermatology 2020. 83, 1323–1330.

8. HIV, P. o. A. T. a. M. M. o. C. L. w. Guidelines for the Use of Antiretroviral Agents in Pediatric HIV Infection in,2022 Department of Health and Human Services, Department of Health and Human Services.

9. Schweitzer, A., Horn, J., Mikolajczyk, R. T., Krause, G. and Ott, J. J. Estimations of worldwide prevalence of chronic hepatitis B virus infection: a systematic review of data published between 1965 and 2013, Lancet 2015. 386, 1546–55.

10. Shepard, C. W., Simard, E. P., Finelli, L., Fiore, A. E. and Bell, B. P. Hepatitis B virus infection: epidemiology and vaccination, Epidemiol Rev 2006. 28, 112–25.

11. Ott, J. J., Stevens, G. A. and Wiersma, S. T. The risk of perinatal hepatitis B virus transmission: hepatitis B e antigen (HBeAg) prevalence estimates for all world regions, BMC Infect Dis 2012. 12, 131.

12. Ott, J. J., Stevens, G. A., Groeger, J. and Wiersma, S. T. Global epidemiology of hepatitis B virus infection: new estimates of age-specific HBsAg seroprevalence and endemicity, Vaccine 2012. 30, 2212–9.

13. McGovern, B. Antiretroviral therapy for patients with HIV-hepatitis B virus coinfection, Clinical infectious diseases : an official publication of the Infectious Diseases Society of America 2007. 44, 1012–3; author reply 1013.

14. Sherman, K. E., Rockstroh, J. and Thomas, D. Human immunodeficiency virus and liver disease: an update in,2015 Wiley Online Library

15. Zeng, D. W., Liu, Y. R., Dong, J., Zhu, Y. Y., Li, Y. B., Chen, J., Zheng, Q. and Jiang, J. J. J. M. m. r. Serum HBsAg and HBeAg levels are associated with liver pathological stages in the immune clearance phase of hepatitis B virus chronic infection, 2015. 11, 3465–3472.

16. Lee, J. H., Hong, S., Im, J. H., Lee, J. S., Baek, J. H. and Kwon, H. Y. Systematic review and meta-analysis of immune response of double dose of hepatitis B vaccination in HIV-infected patients, Vaccine 2020. 38, 3995–4000.

17. Norouzirad, R., Shakurnia, A. H., Assarehzadegan, M.-A., Serajian, A. and Khabazkhoob, M. J. H. m. Serum levels of anti-hepatitis B surface antibody among vaccinated population aged 1 to 18 years in ahvaz city southwest of iran, 2014. 14.

18. Diamant, E. P., Schechter, C., Hodes, D. S. and Peters, V. B. Immunogenicity of hepatitis B vaccine in human immunodeficiency virus-infected children, Pediatr Infect Dis J 1993. 12, 877–8.

19. Launay, O., Rosenberg, A. R., Rey, D., Pouget, N., Michel, M. L., Reynes, J., Neau, D., Raffi, F., Piroth, L., Carrat, F. and Group, A. H. V.-B. Long-term Immune Response to Hepatitis B Virus Vaccination Regimens in Adults With Human Immunodeficiency Virus 1: Secondary Analysis of a Randomized Clinical Trial, JAMA Intern Med 2016. 176, 603–10.

20. Launay, O., van der Vliet, D., Rosenberg, A. R., Michel, M. L., Piroth, L., Rey, D., Colin de Verdiere, N., Slama, L., Martin, K., Lortholary, O., Carrat, F. and Trial, A. H. V.-B. Safety and immunogenicity of 4 intramuscular double doses and 4 intradermal low doses vs standard hepatitis B vaccine regimen in adults with HIV-1: a randomized controlled trial, JAMA 2011. 305, 1432–40.

21. Rey, D., Piroth, L., Wendling, M. J., Miailhes, P., Michel, M. L., Dufour, C., Haour, G., Sogni, P., Rohel, A., Ajana, F., Billaud, E., Molina, J. M., Launay, O., Carrat, F. and group, A. H. B. B. s. Safety and immunogenicity of double-dose versus standard-dose hepatitis B revaccination in non-responding adults with HIV-1 (ANRS HB04 B-BOOST): a multicentre, open-label, randomised controlled trial, Lancet Infect Dis 2015. 15, 1283–91.

22. Chaiklang, K., Wipasa, J., Chaiwarith, R., Praparattanapan, J. and Supparatpinyo, K. Comparison of immunogenicity and safety of four doses and four double doses vs. standard doses of hepatitis B vaccination in HIV-infected adults: a randomized, controlled trial, PLoS One 2013. 8, e80409.

23. Fonseca, M. O., Pang, L. W., de Paula Cavalheiro, N., Barone, A. A. and Heloisa Lopes, M. Randomized trial of recombinant hepatitis B vaccine in HIV-infected adult patients comparing a standard dose to a double dose, Vaccine 2005. 23, 2902–8.

24. Chaiwarith, R., Praparattanapan, J., Kotarathititum, W., Wipasa, J., Chaiklang, K. and Supparatpinyo, K. Higher rate of long-term serologic response of four double doses vs. standard doses of hepatitis B vaccination in HIV-infected adults: 4-year follow-up of a randomised controlled trial, AIDS research and therapy 2019. 16, 33.

25. Laksananun, N., Praparattanapan, J., Kotarathititum, W., Supparatpinyo, K. and Chaiwarith, R. Immunogenicity and safety of 4 vs. 3 standard doses of HBV vaccination in HIV-infected adults with isolated anti-HBc antibody, AIDS research and therapy 2019. 16, 10.

26. Cruciani, M., Mengoli, C., Serpelloni, G., Lanza, A., Gomma, M., Nardi, S., Rimondo, C., Bricolo, F., Consolaro, S., Trevisan, M. and Bosco, O. Serologic response to hepatitis B vaccine with high dose and increasing number of injections in HIV infected adult patients, Vaccine 2009. 27, 17–22.

27. Kerneis, S., Launay, O., Turbelin, C., Batteux, F., Hanslik, T. and Boelle, P. Y. Long-term immune responses to vaccination in HIV-infected patients: a systematic review and meta-analysis, Clinical infectious diseases : an official publication of the Infectious Diseases Society of America 2014. 58, 1130–9.

28. Morell, A., Roth-Wicky, B. and Skvaril, F. Immunoglobulin G subclass restriction of antibodies against hepatitis B surface antigen, Infect Immun 1983. 39, 565–8.

29. Guidotti, L. G., Isogawa, M. and Chisari, F. V. Host-virus interactions in hepatitis B virus infection, Curr Opin Immunol 2015. 36, 61–6.

30. Mosmann, T. R. and Sad, S. The expanding universe of T-cell subsets: Th1, Th2 and more, Immunol Today 1996. 17, 138–46.

31. Urban, M., Winkler, T., Landini, M. P., Britt, W. and Mach, M. Epitope-specific distribution of IgG subclasses against antigenic domains on glycoproteins of human cytomegalovirus, J Infect Dis 1994. 169, 83–90.

32. Tchouangueu, T. F., Lissom, A., Ngu, L. N., NjambePriso, G. D., Tchadji, J. C., Ngane, C. S. S., Ambada, G., Nji, N. N., Ndenkeh, J. J. and Djukouo, L. Vertically Acquired HIV Infection in Children Modulates Hepatitis B Surface Antigen Specific IgG Subclass Distribution After Early Childhood Vaccination Against Hepatitis B Virus Infection, 2018.

33. Tchouangueu, T. F., Mabeku, L. B. K., Lissom, A., Ngu, L. N., Tchuandom, S. B., Tchadji, J. C., Djukouo, L., Ngane, C. S. S., Ngoh, A. A. and Ouambo, H. F. Antibody Responses Specific to Hepatitis B Virus Vaccine in Children Exposed In-Utero to Antiretroviral Therapy, Journal of Clinical & Experimental Immunology 2019. 4.

34. Alter, M. J. Epidemiology of viral hepatitis and HIV co-infection, Journal of Hepatology 2006. 44, S6–S9.

35. Modi, A. A. and Feld, J. J. Viral hepatitis and HIV in Africa, AIDS Reviews 2007. 9, 25–39.

36. Rey-Cuille, M.-A., Njouom, R., Bekondi, C., Seck, A., Gody, C., Bata, P., Garin, B., Maylin, S., Chartier, L., Simon, F. and Vray, M. Hepatitis B Virus Exposure During Childhood in Cameroon, Central African Republic and Senegal After the Integration of HBV Vaccine in the Expanded Program on Immunization, 2013. 32, 1110–1115.

37. Salpini, R., Fokam, J., Ceccarelli, L., Santoro, M. M., Nanfack, A., Sosso, S. M., Kowo, M., Cento, V., Torimiro, J., Sarmati, L., Andreoni, M., Colizzi, V., Perno, C. F. and Njoya, O. High Burden of HBV-Infection and Atypical HBV Strains among HIV-infected Cameroonians, Current HIV research 2016. 14, 165–71.

38. Njom Nlend, A. E., Nguwoh, P. S., Ngounouh, C. T., Tchidjou, H. K., Pieme, C. A., Otele, J. M., Penlap, V., Colizzi, V., Moyou, R. S. and Fokam, J. HIV-Infected or - Exposed Children Exhibit Lower Immunogenicity to Hepatitis B Vaccine in Yaounde, Cameroon: An Appeal for Revised Policies in Tropical Settings?, PLoS One 2016. 11, e0161714.

39. Shaver, Z. M., Anderson, M., Bhebhe, L., Baruti, K., Choga, W. T., Ngidi, J., Mbangiwa, T., Tau, M., Setlhare, D. R., Melamu, P., Phinius, B. B., Musonda, R., Mine, M., Moyo, S. and Gaseitsiwe, S. Decreased hepatitis B virus vaccine response among HIV-positive infants compared with HIV-negative infants in Botswana, AIDS 2022. 36, 755–762.

40. Scolfaro, C., Fiammengo, P., Balbo, L., Madon, E. and Tovo, P. A. Hepatitis B vaccination in HIV-1-infected children: double efficacy doubling the paediatric dose, AIDS 1996. 10, 1169–70.

41. Pessoa, S. D., Miyamoto, M., Ono, E., Gouvea, A. F., de Moraes-Pinto, M. I. and Succi, R. C. Persistence of vaccine immunity against hepatitis B virus and response to revaccination in vertically HIV-infected adolescents on HAART, Vaccine 2010. 28, 1606–12.

42. Lao-araya, M., Puthanakit, T., Aurpibul, L., Sirisanthana, T. and Sirisanthana, V. Antibody response to hepatitis B re-vaccination in HIV-infected children with immune recovery on highly active antiretroviral therapy, Vaccine 2007. 25, 5324–9.

43. Mutwa, P. R., Boer, K. R., Rusine, J. B., Muganga, N., Tuyishimire, D., Reiss, P., Lange, J. M. and Geelen, S. P. Hepatitis B virus prevalence and vaccine response in HIV-infected children and adolescents on combination antiretroviral therapy in Kigali, Rwanda, Pediatr Infect Dis J 2013. 32, 246–51.

44. Obaro, S. K., Pugatch, D. and Luzuriaga, K. Immunogenicity and efficacy of childhood vaccines in HIV-1-infected children, Lancet Infect Dis 2004. 4, 510–8.

45. Beghin, J. C., Ruelle, J., Sokal, E., Bachy, A., Krishna, M., Hall, L., Goubau, P. and Van der Linden, D. Effectiveness of the South African expanded program of immunization against hepatitis B in children infected with human immunodeficiency virus-1 living in a resource-limited setting of Kwazulu-Natal, J Med Virol 2017. 89, 182–185.

46. Fournier, A. M., Baillat, V., Alix-Panabieres, C., Fondere, J. M., Merle, C., Segondy, M., Huguet, M. F., Reynes, J. and Vendrell, J. P. Dynamics of spontaneous HIV-1 specific and non-specific B-cell responses in patients receiving antiretroviral therapy, AIDS 2002. 16, 1755–1760.

47. Nilssen, D. E., Oktedalen, O. and Brandtzaeg, P. Intestinal B cell hyperactivity in AIDS is controlled by highly active antiretroviral therapy, Gut 2004. 53, 487–493.

48. Notermans, D. W., de Jong, J. J., Goudsmit, J., Bakker, M., Roos, M. T., Nijholt, L., Cremers, J., Hellings, J. A., Danner, S. A. and de Ronde, A. Potent antiretroviral therapy initiates normalization of hypergammaglobulinemia and a decline in HIV type 1-specific antibody responses, AIDS Research and Human Retroviruses 2001. 17, 1003–1008.

49. Pensieroso, S., Cagigi, A., Palma, P., Nilsson, A., Capponi, C., Freda, E., Bernardi, S., Thorstensson, R., Chiodi, F. and Rossi, P. Timing of HAART defines the integrity of memory B cells and the longevity of humoral responses in HIV-1 vertically-infected children, Proceedings of the National Academy of Sciences 2009. 106, 7939–7944.

50. Tarlinton, D. B-cell memory: are subsets necessary?, Nature Reviews Immunology 2006. 6, 785–790.

51. Moir, S., Malaspina, A., Ho, J., Wang, W., Dipoto, A. C., O’Shea, M. A., Roby, G., Mican, J. M., Kottilil, S., Chun, T. W., Proschan, M. A. and Fauci, A. S. Normalization of B cell counts and subpopulations after antiretroviral therapy in chronic HIV disease, Journal of Infectious Diseases 2008. 197, 572–579.

52. De Milito, A., Morch, C., Sonnerborg, A. and Chiodi, F. Loss of memory (CD27) B lymphocytes in HIV-1 infection, AIDS 2001. 15, 957–964.

53. D’Orsogna, L. J., Krueger, R. G., McKinnon, E. J. and French, M. A. Circulating memory B-cell subpopulations are affected differently by HIV infection and antiretroviral therapy, AIDS 2007. 21, 1747–1752.

54. Fuster, F., Vargas, J. I., Jensen, D., Sarmiento, V., Acuna, P., Peirano, F., Fuster, F., Arab, J. P., Martinez, F. and Core, H. I. V. S. G. CD4/CD8 ratio as a predictor of the response to HBV vaccination in HIV-positive patients: A prospective cohort study, Vaccine 2016. 34, 1889–1895.

55. Deeks, S. G. HIV infection, inflammation, immunosenescence, and aging, Annual Review of Medicine 2011. 62, 141–155.

56. Serrano-Villar, S., Moreno, S., Fuentes-Ferrer, M., Sanchez-Marcos, C., Avila, M., Sainz, T., de Villar, N. G., Fernandez-Cruz, A. and Estrada, V. The CD4:CD8 ratio is associated with markers of age-associated disease in virally suppressed HIV-infected patients with immunological recovery, HIV Medicice 2014. 15, 40–49.

57. Mussini, C., Lorenzini, P., Cozzi-Lepri, A., Lapadula, G., Marchetti, G., Nicastri, E., Cingolani, A., Lichtner, M., Antinori, A., Gori, A., d’Arminio Monforte, A. and Icona Foundation Study, G. CD4/CD8 ratio normalisation and non-AIDS-related events in individuals with HIV who achieve viral load suppression with antiretroviral therapy: an observational cohort study, Lancet Human Immunodeficiency Virus 2015. 2, e98–106.

58. Moir, S., Malaspina, A., Pickeral, O. K., Donoghue, E. T., Vasquez, J., Miller, N. J., Krishnan, S. R., Planta, M. A., Turney, J. F., Justement, J. S., Kottilil, S., Dybul, M., Mican, J. M., Kovacs, C., Chun, T. W., Birse, C. E. and Fauci, A. S. Decreased survival of B cells of HIV-viremic patients mediated by altered expression of receptors of the TNF superfamily, The Journal of experimental medicine 2004. 200, 587–99.

59. Gougeon, M. L., Lecoeur, H., Dulioust, A., Enouf, M. G., Crouvoiser, M., Goujard, C., Debord, T. and Montagnier, L. Programmed cell death in peripheral lymphocytes from HIV-infected persons: increased susceptibility to apoptosis of CD4 and CD8 T cells correlates with lymphocyte activation and with disease progression, Journal of Immunology 1996. 156, 3509–3520.

60. Ho, J., Moir, S., Malaspina, A., Howell, M. L., Wang, W., DiPoto, A. C., O’Shea, M. A., Roby, G. A., Kwan, R., Mican, J. M., Chun, T. W. and Fauci, A. S. Two overrepresented B cell populations in HIV-infected individuals undergo apoptosis by different mechanisms, Proceedings of the National Academy of Sciences 2006. 103, 19436–19441.

61. De Milito, A., Nilsson, A., Titanji, K., Thorstensson, R., Reizenstein, E., Narita, M., Grutzmeier, S., Sonnerborg, A. and Chiodi, F. Mechanisms of hypergammaglobulinemia and impaired antigen-specific humoral immunity in HIV-1 infection, Blood 2004. 103, 2180–2186.

62. Hart, M., Steel, A., Clark, S. A., Moyle, G., Nelson, M., Henderson, D. C., Wilson, R., Gotch, F., Gazzard, B. and Kelleher, P. Loss of discrete memory B cell subsets is associated with impaired immunization responses in HIV-1 infection and may be a risk factor for invasive pneumococcal disease, Journal of Immunology 2007. 178, 8212–8220.

63. Titanji, K., De Milito, A., Cagigi, A., Thorstensson, R., Grutzmeier, S., Atlas, A., Hejdeman, B., Kroon, F. P., Lopalco, L., Nilsson, A. and Chiodi, F. Loss of memory B cells impairs maintenance of long-term serologic memory during HIV-1 infection, Blood 2006. 108, 1580–1587.

64. Malaspina, A., Moir, S., Orsega, S. M., Vasquez, J., Miller, N. J., Donoghue, E. T., Kottilil, S., Gezmu, M., Follmann, D., Vodeiko, G. M., Levandowski, R. A., Mican, J. M. and Fauci, A. S. Compromised B cell responses to influenza vaccination in HIV-infected individuals, Journal of Infectious Diseases 2005. 191, 1442–1450.

65. Wawer, M. J., Gray, R. H., Sewankambo, N. K., Serwadda, D., Paxton, L., Berkley, S., McNairn, D., Wabwire-Mangen, F., Li, C., Nalugoda, F., Kiwanuka, N., Lutalo, T., Brookmeyer, R., Kelly, R. and Quinn, T. C. A randomized, community trial of intensive sexually transmitted disease control for AIDS prevention, Rakai, Uganda, AIDS 1998. 12, 1211–1225.

66. Gray, R. H., Wawer, M. J., Serwadda, D., Sewankambo, N., Li, C., Wabwire-Mangen, F., Paxton, L., Kiwanuka, N., Kigozi, G., Konde-Lule, J., Quinn, T. C., Gaydos, C. A. and McNairn, D. Population-based study of fertility in women with HIV-1 infection in Uganda, Lancet 1998. 351, 98–103.

67. Homsy, J., Bunnell, R., Moore, D., King, R., Malamba, S., Nakityo, R., Glidden, D., Tappero, J. and Mermin, J. Reproductive intentions and outcomes among women on antiretroviral therapy in rural Uganda: a prospective cohort study, PloS One 2009. 4, e4149.

68. van Leth, F., Phanuphak, P., Stroes, E., Gazzard, B., Cahn, P., Raffi, F., Wood, R., Bloch, M., Katlama, C., Kastelein, J. J., Schechter, M., Murphy, R. L., Horban, A., Hall, D. B., Lange, J. M. and Reiss, P. Nevirapine and efavirenz elicit different changes in lipid profiles in antiretroviral-therapy-naive patients infected with HIV-1, PLoS Med 2004. 1, e19.

69. van Leth, F., Phanuphak, P., Ruxrungtham, K., Baraldi, E., Miller, S., Gazzard, B., Cahn, P., Lalloo, U. G., van der Westhuizen, I. P., Malan, D. R., Johnson, M. A., Santos, B. R., Mulcahy, F., Wood, R., Levi, G. C., Reboredo, G., Squires, K., Cassetti, I., Petit, D., Raffi, F., Katlama, C., Murphy, R. L., Horban, A., Dam, J. P., Hassink, E., van Leeuwen, R., Robinson, P., Wit, F. W., Lange, J. M. and team, N. N. S. Comparison of first-line antiretroviral therapy with regimens including nevirapine, efavirenz, or both drugs, plus stavudine and lamivudine: a randomised open-label trial, the 2NN Study, Lancet 2004. 363, 1253–1263.

70. Ford, N. and Calmy, A. Improving first-line antiretroviral therapy in resource-limited settings, Curr Opin HIV AIDS 2010. 5, 38–47.

71. Ford, N., Mofenson, L., Kranzer, K., Medu, L., Frigati, L., Mills, E. J. and Calmy, A. Safety of efavirenz in first-trimester of pregnancy: a systematic review and meta-analysis of outcomes from observational cohorts, AIDS 2010. 24, 1461–1470.

72. Myer, L., Carter, R. J., Katyal, M., Toro, P., El-Sadr, W. M. and Abrams, E. J. Impact of antiretroviral therapy on incidence of pregnancy among HIV-infected women in Sub-Saharan Africa: a cohort study, PLoS Med 2010. 7, e1000229.

