## Supplementary figures and images for "Could Anti-retroviral treatment affects the profile of hepatitis B vaccine specific antibody in vertically HIV-infected children?"

### Supplemental Figure 1

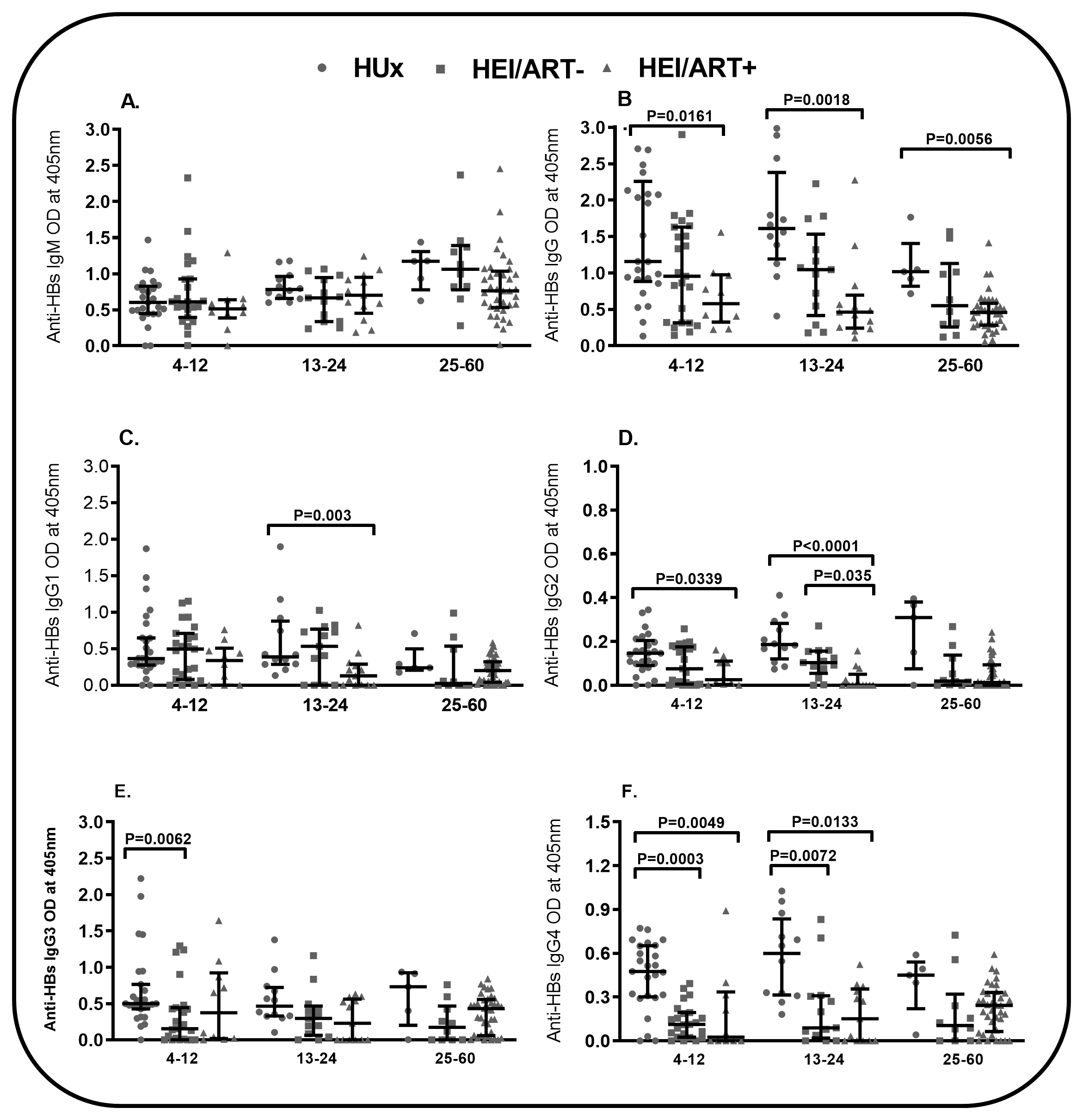
